# Temporal Deep Learning for Predicting Periodontitis Progression Using Longitudinal Gingival Crevicular Fluid Protein Profiles

**DOI:** 10.64898/2026.03.11.26348163

**Authors:** Zoe Xiaofang Zhu, Jake Chen, Flavia Teles

## Abstract

**Background:** Conventional clinical indicators of periodontitis progression detect disease after irreversible tissue destruction has occurred. Molecular biomarkers in gingival crevicular fluid (GCF) offer potential for earlier detection, but existing analytical approaches rely on cross-sectional snapshots that fail to capture the temporal dynamics of disease evolution.

**Aim:** To develop and validate a temporal deep learning framework leveraging longitudinal GCF protein profiles for (1) regression-based prediction of clinical attachment level (CAL) and probing depth (PD) changes, (2) current-visit classification of periodontitis progression, (3) next-visit prediction of progression with a 2-month clinical lead time, and (4) identification of the most informative biomarkers through systematic multi-method feature importance analysis.

**Materials and Methods:** This study utilized longitudinal GCF data from a prospective cohort of 413 participants (501 periodontal sites, 3,792 time-series observations) with 64 protein biomarkers measured at 2-month intervals over 12 months. A compact encoder-gated recurrent unit (GRU)-decoder architecture was developed through systematic experimentation across four phases, benchmarking temporal deep learning against cross-sectional machine learning baselines. Task-specific decoders addressed continuous regression (CAL and PD prediction) and binary classification (progression detection). Model development and reporting followed the TRIPOD+AI guidelines.

**Results:** The temporal GRU achieved 47.7% CAL mean absolute error (MAE) reduction (1.139 to 0.596 mm) and 41.0% PD MAE reduction (0.902 to 0.532 mm) over linear regression baselines through the systematic model development progression. For binary classification, the model achieved AUC-ROC of 0.886 for current-visit classification and 0.867 for next-visit prediction with a 2-month lead time. Per-visit analysis revealed progressive improvement in both regression and classification accuracy as longitudinal data accumulated. Cross-method feature importance analysis identified Periostin, VEGF, MMP-2, IL-1RA, and MCP-4 as core predictive biomarkers, with divergent profiles between diagnostic and prognostic tasks suggesting distinct molecular signatures for concurrent versus incipient progression.

**Conclusions:** Temporal deep learning applied to longitudinal GCF protein profiles enables both accurate regression prediction of clinical parameters and reliable classification of progression status, including 2-month-ahead forecasting suitable for clinical intervention planning. The compact architecture and non-invasive sampling approach make this framework suitable for integration into point-of-care periodontal monitoring workflows.

**Clinical Relevance:** Conventional clinical indicators of periodontitis progression, including probing depth changes, attachment loss, and radiographic bone loss, inherently detect disease after irreversible damage has occurred. This study shows that a compact deep learning model analyzing temporal GCF protein profiles can first accurately predict continuous changes in pocket depth and attachment loss, then classify progression status 2 months in advance, enabling proactive intervention before clinical manifestation of tissue destruction.

## 1. Introduction

Periodontitis is a chronic, multifactorial inflammatory disease that results in progressive destruction of the tooth-supporting apparatus and represents a leading cause of tooth loss globally^1^. Affecting an estimated 1.1 billion individuals worldwide in its severe form^2^, periodontitis imposes substantial healthcare costs and has systemic implications linked to cardiovascular disease, diabetes, and adverse pregnancy outcomes^3^. Despite substantial advances in understanding its pathobiology, clinical practice remains fundamentally reactive. The standard diagnostic armamentarium (including probing pocket depth, clinical attachment level (CAL), bleeding on probing, and radiographic assessment) can only confirm tissue destruction that has already occurred^4^. This inherent diagnostic lag means that therapeutic intervention is frequently initiated after significant and irreversible structural compromise, limiting the opportunity for disease interception.

Gingival crevicular fluid (GCF) offers a molecularly rich and site-specific window into the biological processes underlying periodontal tissue homeostasis and breakdown. As an inflammatory exudate derived from the gingival sulcus, GCF contains a complex proteomic signature that reflects real-time host immune responses, microbial challenge, and tissue remodeling activity at individual periodontal sites^5^. In contrast to systemic biomarkers in serum or whole saliva, the site-specificity of GCF makes it particularly valuable for monitoring localized disease activity. Previous investigations have identified associations between individual GCF biomarkers and periodontal disease severity, including matrix metalloproteinases, particularly MMP-8^6^, pro-inflammatory cytokines such as IL-1beta and IL-6^7^, and bone metabolism mediators including RANKL and osteoprotegerin^8^. However, single-biomarker approaches have generally shown limited predictive capacity for disease progression, as the pathobiological complexity of periodontitis involves coordinated alterations across multiple molecular pathways simultaneously.

Recent work has demonstrated the superiority of multi-biomarker panel approaches over single-marker assays. Grant et al. identified a panel of GCF and salivary proteins, including MMP-9, S100A8, and alpha-1-acid glycoprotein, that effectively discriminated between periodontal health and disease^9^. Huang et al. developed a periodontal disease antibody array targeting 20 disease-related proteins and compared five machine learning classifiers, achieving strong classification accuracy with a panel anchored by IL-1β, IL-8, and MMP-13^10^. In the parent cohort of the present study, longitudinal analysis of salivary and serum inflammatory biomarkers established the temporal dynamics of systemic and local inflammatory responses during disease progression and after treatment^11^. Nevertheless, these prior approaches relied on cross-sectional feature extraction or simple tabular representations of longitudinal data, without explicitly modeling the temporal dynamics of biomarker trajectories.

The application of artificial intelligence in periodontology has grown rapidly, though predominantly focused on image-based approaches. Convolutional neural networks have shown promise for automated detection of radiographic bone loss^12^, while machine learning classifiers using clinical and radiographic features have achieved strong performance for disease staging^13,14^. However, these approaches operate on static snapshots rather than temporal sequences, and none have leveraged the rich longitudinal molecular information available from serial GCF sampling.

Temporal deep learning architectures, particularly recurrent neural networks (RNNs) such as gated recurrent units (GRUs) and long short-term memory (LSTM) networks^15,16^, are specifically designed to capture sequential dependencies in time-series data. In clinical medicine, these architectures have demonstrated strong performance for disease trajectory prediction from electronic health records^17,18^, ICU outcome forecasting^19^, and medication response prediction^20^. Attention mechanisms further enhance these architectures by learning to weight the relative importance of different timepoints, enabling the model to focus on critical disease trajectory inflection points^17,21^. Despite this proven capability for temporal clinical data, recurrent neural network approaches that explicitly model the longitudinal evolution of molecular biomarker profiles remain entirely unexplored in periodontology.

The aims of this study were fourfold: (1) to develop a temporal deep learning framework for regression-based prediction of CAL and PD changes from longitudinal GCF protein profiles, validating the superiority of temporal over cross-sectional modeling through systematic experimentation; (2) to evaluate current-visit classification of periodontitis progression status using the same architectural framework; (3) to determine whether this framework can predict progression at the next clinical visit, providing a clinically actionable 2-month intervention window; and (4) to identify the most informative GCF biomarkers through systematic multi-method feature importance analysis, providing biological insight and a foundation for targeted biomarker panels.

## 2. Materials and Methods

### 2.1 Study design and reporting guidelines

This study is a secondary analysis of data from a prospective longitudinal cohort of periodontitis patients. Model development and reporting followed the TRIPOD+AI guidelines for clinical prediction model studies^22^. The study protocol was approved by the Institutional Review Board of the Tufts University (IRB ID: STUDY00006807).

### 2.2 Study population and dataset

The parent cohort, study design, participant recruitment, eligibility criteria, clinical examination procedures, and demographic characteristics have been described in detail previously^11,23^. Briefly, the cohort comprised 413 participants with chronic periodontitis who were monitored at 2-month intervals over a 12-month observation period (visit months 0, 2, 4, 6, 8, 10, and 12). A total of 501 periodontal sites were selected for longitudinal monitoring based on baseline disease severity criteria. At each visit, GCF samples were collected from the monitored sites using standardized paper strip absorption, and clinical parameters including CAL and probing pocket depth (PD) were recorded by calibrated examiners.

### 2.3 GCF biomarker quantification

At each visit, 64 GCF protein biomarkers were quantified using commercially available Milliplex immunoassay panels (Millipore, Burlington, MA, USA). The biomarker panel encompassed six functional categories: cytokines involved in inflammatory and immune regulation (IFN-γ, IFN-α2, IL-1α, IL-1β, IL-2, IL-6, IL-7, IL-8, IL-10, IL-12p40, IL-12p70, IL-13, IL-15, IL-16, IL-17A, IL-21, IL-23, IL-28A, TNF-α, IL-1RA), matrix metalloproteinases (MMP-1, MMP-2, MMP-8, MMP-9, MMP-10), chemokines and chemoattractants (MCP-1, MCP-3, MCP-4, MIP-1α, MIP-1β, MIP-1δ, Eotaxin, Eotaxin-2, I-309/CCL1, IP-10, RANTES, ENA-78, BCA-1, MDC, Fractalkine, SDF-1α+β, 6Ckine, GRO), growth factors and hematopoietic cytokines (VEGF, EGF, TGF-α, PDGF-AA, G-CSF, GM-CSF, Flt-3L, SCF, LIF), bone and tissue remodeling markers (Periostin, DKK1, OPG, RANKL, TRAP5, YKL-40), immune activation and cytotoxic mediators (TSLP, TRAIL, sCD40L, Granzyme B), and stress-response proteins (HSP70, GDF-15).This panel was selected to provide broad coverage of the molecular pathways implicated in periodontal tissue homeostasis and destruction, including innate and adaptive immunity, extracellular matrix turnover, angiogenesis, bone remodeling, and stress responses.

### 2.4 Data preprocessing and missing data handling

All biomarker concentrations were log-transformed (log10) prior to analysis to normalize the typically right-skewed distributions of protein concentration data. Of the original 3,808 observations collected across all participants and visits, 16 samples (0.4%) with missing biomarker data were excluded from the analysis without imputation, yielding 3,792 complete time-series observations. This exclusion rate was considered negligible and unlikely to introduce systematic bias. For the temporal models, variable-length sequences arising from missed visits were handled using binary masks and a sentinel value of −1.0, allowing the model to distinguish between true biomarker measurements and missing timepoints without introducing artificial values.

### 2.5 Progression definition

Periodontitis progression was defined as a CAL increase of 2 mm or more within each 2-month observation interval, consistent with established clinical thresholds for meaningful attachment loss at the site level^24^ and aligned with the 2017 World Workshop framework for characterizing disease progression rates^25,26^. Binary progression labels were assigned for each interval. The baseline visit (month 0) was excluded from classification targets, serving as the temporal reference.

### 2.6 Data partitioning

Stratified random splitting by site-level progression status yielded a training set (n = 289), a validation set (n = 62), and a held-out test set (n = 62), following an approximate 70/15/15 split. All longitudinal observations from a given participant were confined to a single partition. Temporal sequences were constructed by grouping observations by subject-site pairs and ordering by visit month, with binary masks applied to handle variable sequence lengths.

### 2.7 Systematic model development

The model architecture was developed through a systematic experimental progression across four modeling phases (Figure 1B). Phase 1 established cross-sectional machine learning baselines (linear regression, random forest, XGBoost) using single-visit biomarker snapshots, providing a floor for expected performance from non-temporal approaches. Phase 2 evaluated a multi-layer perceptron encoder to assess the benefit of learned non-linear feature representations without temporal modeling. Phase 3 introduced the GRU temporal backbone, enabling the explicit modeling of sequential biomarker dynamics across visits. Phase 4 systematically evaluated architectural variants including sinusoidal time encoding, multiple attention mechanisms (additive Bahdanau-style^27^, multiplicative Luong-style^28^, weighted-sum sigmoid^29^), and incremental prediction strategies (predicting delta change rather than absolute values). Model selection at each phase was guided by regression MAE on the validation set, with the best-performing variant carried forward.

**FIGURE 1.**
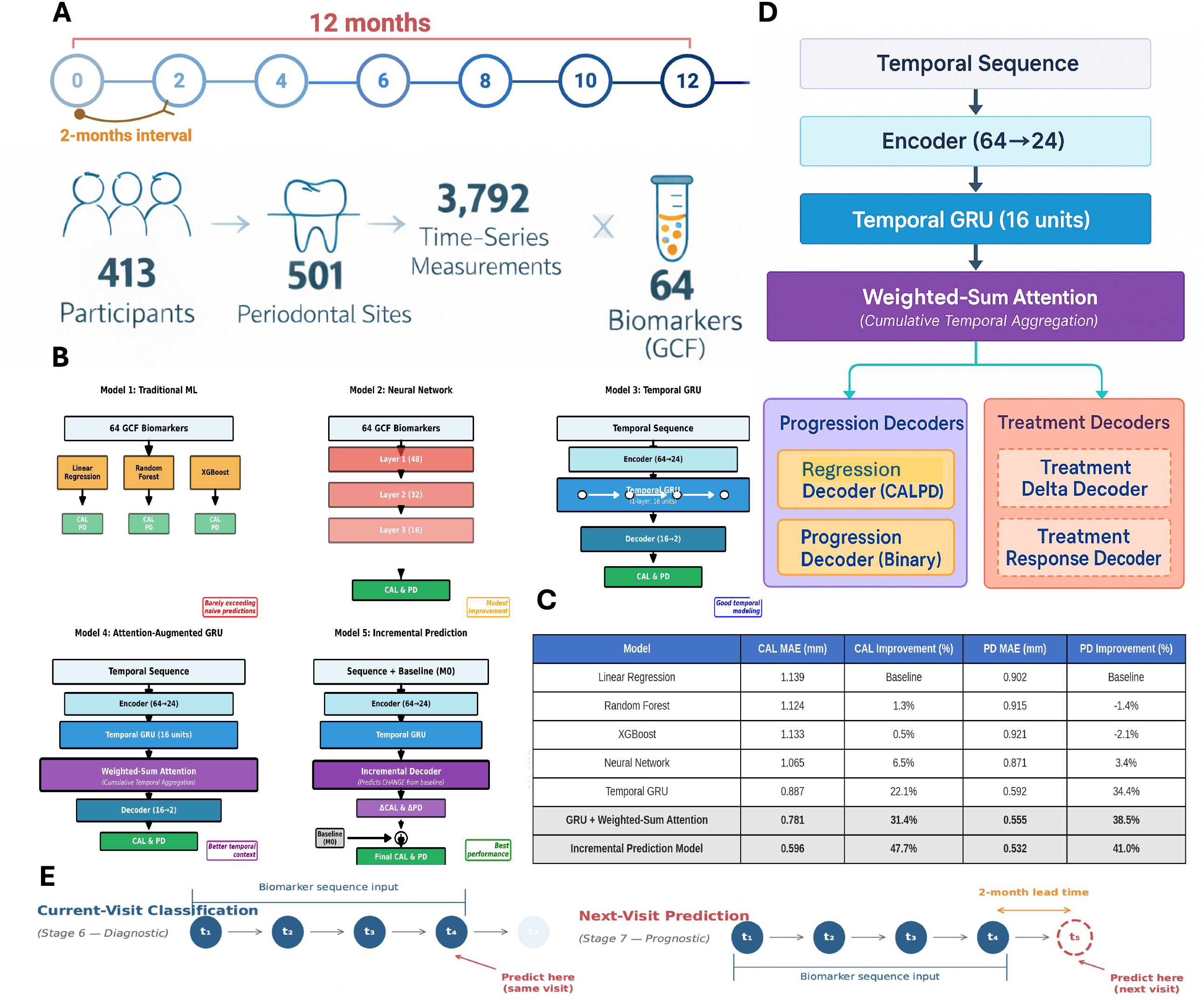

### 2.8 Final model architecture

The Phase 3 GRU-based architecture, further refined through Phase 4 experimentation, yielded the final encoder–GRU–decoder design (Figure 1D). The biomarker encoder compresses the 64-dimensional input at each timepoint to a lower-dimensional latent representation through linear layers with Rectified Linear Unit (ReLU) activation and dropout regularization, reducing dimensionality while preserving inter-biomarker relationships. The temporal module consists of a single-layer unidirectional GRU with 16 hidden units that processes the encoded sequence causally, learning to extract disease trajectory patterns from the ordered biomarker observations across visits. The GRU was selected over LSTM due to its comparable performance with fewer parameters, which is advantageous given the limited training set size.

Two task-specific decoders branch from this temporal backbone, reflecting the complementary nature of the prediction objectives. The regression decoder maps the GRU hidden state through an intermediate layer to two continuous outputs representing predicted CAL and PD values. The progression classification decoder maps the hidden state directly to a single logit indicating progression probability. This design reflects the conceptual relationship between the tasks: periodontitis progression is defined by changes in CAL, such that the regression task estimates the continuous clinical measure underlying the binary classification outcome.

An attention-augmented variant was additionally evaluated for the binary classification tasks. This variant incorporates a weighted-sum attention mechanism with sigmoid-based cumulative aggregation over temporal hidden states, implemented as a two-layer scoring network (16 → 16 → 1 with Tanh activation) that learns visit-specific importance weights. The causal aggregation ensures that predictions at each timepoint attend only to current and preceding visits, preserving temporal validity and preventing information leakage from future observations.

### 2.9 Training procedure

Models were trained using task-appropriate loss functions: L1 loss (mean absolute error) for regression and binary cross-entropy loss with logits (BCEWithLogitsLoss) for classification. The classification loss incorporated automatically computed positive class weighting to address class imbalance. This weighting effectively upsamples the positive class in the loss computation, ensuring that rare progression events contribute meaningfully to gradient updates despite their low prevalence. Optimization was performed using the AdamW optimizer with weight decay regularization and ReduceLROnPlateau learning rate scheduling. Training continued for up to 200 epochs with early stopping on validation loss to prevent overfitting. All models were implemented in PyTorch Lightning with a batch size of 32, and training completed in under 5 minutes per variant on a single GPU.

### 2.10 Regression prediction task

The regression task evaluates the temporal model’s capacity to predict continuous CAL and PD values at each visit using the accumulated biomarker sequence. This task served as the primary optimization target during the systematic model development phases (Phases 1 through 4), as continuous prediction provides richer gradient information for architecture selection and hyperparameter tuning than binary classification. Mean absolute error (MAE) was computed separately for CAL and PD predictions on the held-out test set, serving as the development metric. Per-visit MAE analysis assessed how prediction accuracy evolves as longitudinal evidence accumulates, providing insight into the minimum monitoring duration required for reliable predictions.

### 2.11 Binary classification tasks

Two clinically motivated binary classification tasks were evaluated (Figure 1E). For current status classification (diagnostic), the model predicts the progression status at the same visit where biomarkers were measured, using the accumulated temporal sequence up to and including that visit. For future progression prediction (prognostic), the model predicts the progression status at the subsequent visit (t + 1) using the biomarker sequence through the current visit (t), providing a 2-month clinical lead time for intervention planning. The primary evaluation metric was AUC-ROC, which is threshold-independent and robust to class imbalance. Secondary metrics included sensitivity, specificity, balanced accuracy, precision, and F1-score. Threshold optimization was performed by evaluating the full range of decision thresholds on the test set to identify operating points for broad population screening, confirmatory assessment, and severe case monitoring. The positive likelihood ratio (LR+) was computed at key operating points to assess clinical utility.

### 2.12 Feature importance analysis

Biomarker importance was assessed through two complementary approaches to ensure robustness of findings. Weight-based importance quantified each biomarker’s contribution via the encoder’s first-layer weight magnitudes using a composite score. This approach captures how strongly the model’s learned representation responds to each input biomarker. Gradient-based importance computed the mean absolute input gradient across all test samples, measuring how much small changes in each biomarker affect the model’s predictions, an approach related to saliency mapping in neural network interpretability^30^. Both methods were applied independently to both the current-visit and next-visit models. Cross-task overlap was assessed to identify core biomarkers, defined as those consistently appearing in the top 10 across both tasks and both methods.

## 3. Results

### 3.1 Systematic model development and regression performance

The systematic experimental progression validated the critical role of temporal modeling for predicting clinical parameter changes (Figure 1C). Phase 1 cross-sectional baselines achieved CAL MAE of 1.124 to 1.139 mm and PD MAE of 0.902 to 0.921 mm, on the held-out test set. These results demonstrate that single-visit biomarker snapshots provide limited predictive capacity for longitudinal clinical changes. Notably, the random forest and XGBoost models showed severe overfitting (training R^2^ > 0.70 vs. test R^2^ < 0.12 for CAL), highlighting the challenge of the limited sample size for non-temporal approaches. The Phase 2 multi-layer perceptron improved to CAL MAE 1.065 mm (6.5% improvement), confirming the ceiling of non-temporal learned representations.

Introduction of the Phase 3 GRU temporal backbone marked a qualitative leap in prediction accuracy, reducing CAL MAE to 0.887 mm (22.1% improvement over baseline) and PD MAE to 0.592 mm (34.4% improvement) with 30% fewer parameters than the MLP (3,882 vs. 5,544). This dramatic improvement upon introducing temporal processing provides strong evidence that the trajectory of biomarker changes over time, rather than their absolute concentrations at any single visit, carries essential prognostic information for periodontal disease progression.

The Phase 4 weighted-sum attention mechanism provided further improvement to CAL MAE 0.781 mm (31.4% cumulative improvement) and PD MAE 0.555 mm (38.5%). The final incremental prediction variant, which predicts change relative to baseline rather than absolute values, achieved the best regression performance: CAL MAE 0.596 mm (47.7% cumulative improvement) and PD MAE 0.532 mm (41.0%). This improvement suggests that modeling relative change removes inter-individual variability in baseline clinical measurements, allowing the model to focus on the dynamics of disease progression independent of each patient’s starting point.

### 3.2 Current-status classification

Using only the 64 GCF protein markers, independently from clinical examination parameters, the current-status classification model achieved an AUC-ROC of 0.886 on the held-out test set (Figure 2A). At the default decision threshold of 0.5, the baseline model (without attention) attained 82.4% sensitivity and 79.6% specificity. The attention-augmented variant achieved higher sensitivity (94.1%) at the expense of specificity (72.9%), yielding an AUC-ROC of 0.863.

**FIGURE 2.**
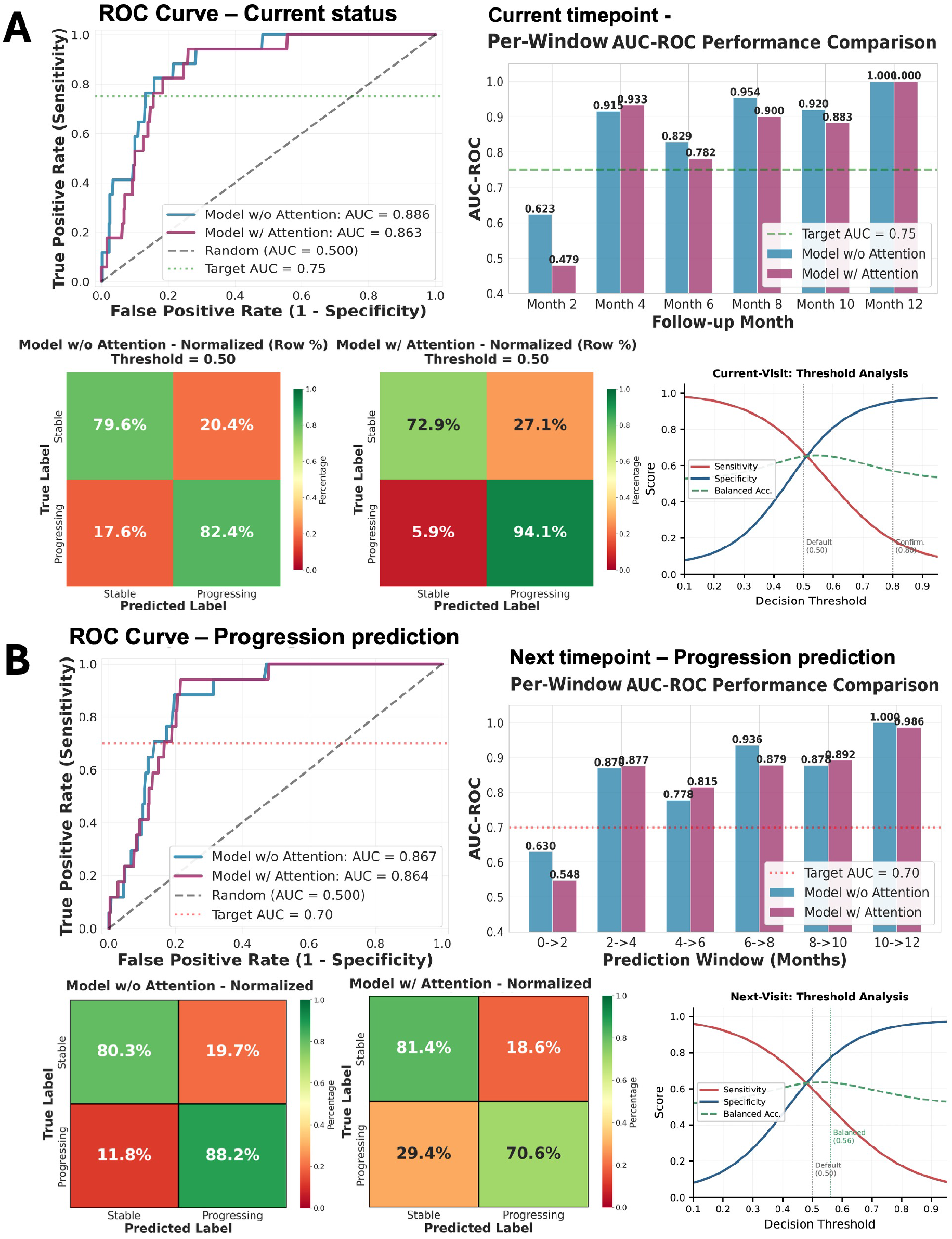

Performance varied systematically across the monitoring period, reflecting the influence of accumulated temporal context. The earliest interval (month 2) showed the lowest discrimination (AUC-ROC 0.623), attributable to the limited temporal context from a single observation. Performance improved progressively as longitudinal data accumulated, with months 4, 8, and 10 achieving AUC-ROC values of 0.916, 0.954, and 0.920, respectively. This pattern mirrors the regression findings and reinforces the importance of serial monitoring for reliable disease assessment.

Threshold optimization identified clinically relevant operating points. A screening threshold of 0.50 maximized sensitivity to 94.1% with specificity of 72.9%, suitable for initial risk assessment where the cost of missing a progression event outweighs the cost of false positives. A confirmatory operating point at 0.80 provided 96.5% specificity and a positive likelihood ratio (LR+) of 11.1, suitable for informing treatment planning decisions when high diagnostic certainty is required. These two thresholds enable a two-stage clinical protocol: broad screening followed by targeted confirmation.

### 3.3 Next-visit progression prediction

The next-visit prediction model demonstrated that GCF biomarker trajectories can forecast progression 2 months before clinical manifestation (Figure 2B). The baseline model achieved an AUC-ROC of 0.867 with 88.2% sensitivity and 80.3% specificity at the default threshold, exceeding the a priori performance target of 0.70 by 23.9%. The attention variant achieved similar AUC-ROC (0.864) but with lower sensitivity (70.6%) and higher specificity (81.4%).

Prediction performance by temporal window revealed clinically informative patterns. The initial 0 to 2 month window showed the lowest performance (AUC-ROC 0.630), as the model must predict future progression using only a single baseline measurement. Beyond this initial window, all subsequent intervals achieved strong discrimination (average AUC-ROC 0.89), confirming that even minimal longitudinal context, as few as two observations, substantially improves predictive accuracy.

The comparable performance between current-visit classification and next-visit prediction is notable, as predicting future events is inherently more challenging than classifying concurrent status. It suggests that molecular precursors of progression are detectable in GCF protein profiles before clinical manifestation, consistent with the biological expectation that inflammatory and tissue remodeling processes precede measurable attachment loss.

### 3.4 Feature importance

Cross-method feature importance analysis revealed a consistent set of core biomarkers driving both classification and prediction (Figure 3A-C). Periostin emerged as the single most important biomarker across analyses, ranking first in weight-based importance for both current-visit (composite score 0.849) and next-visit (0.933) models, and first in gradient-based importance for the next-visit model (score 0.141). VEGF was the second most consistently important marker, appearing in the top three across both weight-based and gradient-based analyses for both tasks. MMP-2 ranked second for current-visit classification by weight-based analysis (score 0.821), while MMP-9 emerged prominently for next-visit prediction (weight rank 9, gradient rank 3), suggesting differential roles of these gelatinases in concurrent versus incipient progression. IL-1RA showed strong importance in both tasks (weight-based rank 8 and 3 for current-visit and next-visit, respectively), while MCP-4 and TSLP completed the core biomarker set appearing consistently across both tasks and methods.

**FIGURE 3.**
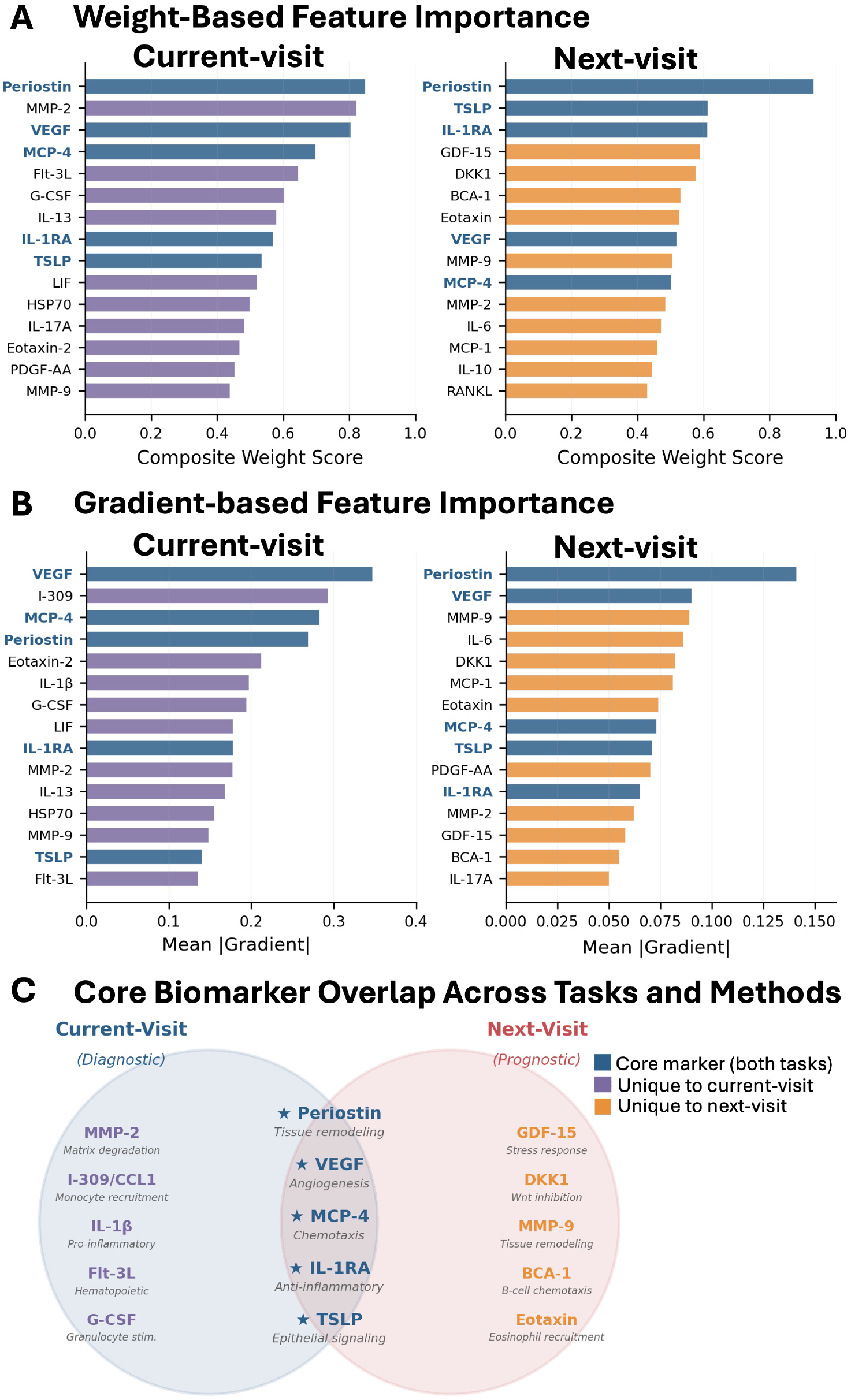

The next-visit model placed greater weight on GDF-15 and DKK1, the markers associated with cellular stress and Wnt signaling inhibition, while gradient-based analysis for the current-visit model highlighted IL-1beta and I-309/CCL1, canonical mediators of active periodontal inflammation. This divergence between diagnostic and predictive biomarker profiles suggests that distinct molecular signatures characterize concurrent active destruction versus the early molecular events that precede clinically detectable progression.

## 4. Discussion

This study presents a temporal deep learning framework for predicting periodontitis progression using longitudinal GCF protein profiles, representing the first application of recurrent neural networks to molecular biomarker time-series data in periodontology. The principal findings are that (1) the temporal GRU architecture achieves a 47.7% improvement in regression MAE over cross-sectional baselines for predicting CAL and PD changes, (2) this same architectural framework enables strong current-visit classification (AUC-ROC 0.886) and 2-month-ahead progression prediction (AUC-ROC 0.867, sensitivity 88.2%), and (3) multi-method feature importance analysis identifies a biologically coherent core biomarker set anchored by Periostin, VEGF, and MMP-2.

### 4.1 The critical role of temporal modeling

The critical contribution of temporal modeling was demonstrated through the systematic regression-based model development. Cross-sectional machine learning baselines using single-visit biomarker profiles yielded negligible improvement over naive prediction. This finding is consistent with the recognized limitations of single-timepoint diagnostic approaches in periodontology^4^ and suggests that the cross-sectional biomarker snapshot captures insufficient information about the dynamic process of disease progression.

The GRU temporal backbone reduced CAL MAE to 0.887 mm with 30% fewer parameters than the MLP. This simultaneous improvement in both accuracy and parameter efficiency is notable: the temporal inductive bias of the GRU serves as an effective regularizer, reducing overfitting while enabling the model to leverage the sequential structure of the data. This finding confirms that the trajectory of biomarker changes over time, rather than their absolute concentrations at any single visit, carries essential prognostic information. The progressive improvement in per-visit regression accuracy further validates that accumulated longitudinal evidence provides increasingly reliable clinical predictions. The incremental prediction variant’s additional 25.6% MAE reduction (from 0.887 to 0.596 mm for CAL). This strategy likely succeeds because it reduces inter-individual variability in baseline clinical measurements. By modeling the change trajectory rather than the absolute level, the model can focus on the dynamics of disease progression independent of each patient’s starting point.

### 4.2 From regression to binary classification

The transition from regression to binary classification demonstrated the clinical utility of the temporal framework. Progression is inherently linked to the regression predictions: a model that accurately estimates continuous CAL changes can in principle classify progression by thresholding its continuous output. Training dedicated binary classification models with weighted loss functions proved more effective for the class-imbalanced clinical setting, as the binary cross-entropy loss with class weighting directly optimizes for the discriminative boundary between progression and stable states, while the regression loss optimizes for continuous accuracy across the entire range of clinical values.

### 4.3 Temporal windows and clinical implementation

The per-window analysis for the next timepoint progression prediction revealed that the 2 to 4 month window already achieved strong discrimination (AUC-ROC 0.878), demonstrating that as few as three sequential observations are sufficient for accurate progression forecasting. This finding has important clinical implications: it suggests that the molecular precursors of periodontal destruction become detectable in GCF proteins within the first few months of monitoring, and that early serial sampling provides the greatest informational gain per visit. Performance continued to strengthen with data accumulation, supporting a clinical protocol of serial GCF sampling with progressive risk refinement.

The threshold optimization analysis provides a practical framework for clinical deployment across different use cases. For broad population screening during routine maintenance visits, a sensitive threshold minimizes missed progression events. For confirmatory assessment where diagnostic certainty is required to guide treatment planning, a specific threshold reduces false positives. For severe case monitoring in high-risk patients, clinicians may prioritize sensitivity to ensure no progression event goes undetected. These thresholds can be applied independently based on the clinical context, or combined in a two-stage screen-then-confirm protocol to balance detection sensitivity with diagnostic precision.

### 4.4 Biological interpretation of feature importance

Feature importance analysis converged on a biologically coherent core biomarker set. Periostin, a matricellular protein essential for periodontal ligament integrity, collagen fibrillogenesis, and wound healing^31^, emerged as the top predictor in nearly all analyses. This finding is consistent with Teles et al. who reported elevated Periostin at stable periodontal sites, suggesting a tissue-protective role^7^. The model’s reliance on Periostin likely reflects its value as an indicator of the balance between tissue destruction and repair processes, reduced Periostin may signal inadequate repair capacity preceding clinical progression.

VEGF (vascular endothelial growth factor) and MMP-2 (gelatinase A) reflect the angiogenic and proteolytic processes underlying progressive attachment loss. VEGF is upregulated in inflamed periodontal tissues and promotes vascular permeability and inflammatory cell recruitment^32^. MMP-2, a gelatinase capable of degrading basement membrane collagen, plays a central role in extracellular matrix remodeling during periodontal tissue destruction^33^. The identification of IL-1RA (interleukin-1 receptor antagonist) underscores the role of endogenous anti-inflammatory responses, as this natural inhibitor of IL-1 signaling may serve as a marker of the host’s capacity to mount protective anti-inflammatory responses.

The divergence between diagnostic and predictive biomarker profiles is particularly informative. The next-visit model placed greater weight on GDF-15 (growth differentiation factor 15, associated with cellular stress responses and mitochondrial dysfunction) and DKK1 (Dickkopf-1, an inhibitor of Wnt signaling involved in bone remodeling), while gradient-based analysis for the current-visit model highlighted IL-1beta and I-309/CCL1 (canonical mediators of active periodontal inflammation and monocyte chemotaxis). This pattern suggests that molecular events preceding progression, including cellular stress and disrupted bone homeostasis, differ from those concurrent with active destruction, which are characterized by acute inflammation and tissue breakdown. Understanding these distinct molecular phases could inform the development of phase-specific therapeutic interventions.

### 4.5 Comparison with existing approaches

Our results compare favorably with recent approaches while offering capabilities not available in existing methods. Furquim et al.^34^ achieved AUC-ROC of 0.88 using a probabilistic graphical model that combined clinical parameters with 10 salivary analytes, and Deng et al.^14^ developed a machine learning screening tool achieving AUC greater than 0.94 for periodontal disease staging using clinical parameters and salivary aMMP-8. The present study achieves comparable current-visit performance using only GCF biomarker profiles without any clinical examination data, suggesting that temporal modeling of molecular signatures can capture prognostic information equivalent to that provided by clinical parameters. This independence from clinical measurements is a key distinction, as it enables potential application in remote monitoring scenarios between routine office visits, a capability not possible with approaches that require concurrent clinical examination. Furthermore, both prior studies addressed cross-sectional classification of current disease status, whereas our framework extends to longitudinal regression prediction and 2-month-ahead temporal forecasting of future progression events.

### 4.6 Limitations

Several limitations merit consideration. First, there is a severe class imbalance with a low positive rate. While this reflects the true epidemiology of site-level progression in a maintenance population, the high false positive rate would generate unnecessary clinical alerts. Prospective validation in cohorts with varying prevalence rates is essential to establish the clinical utility of threshold-based classification.

Second, the dataset derives from a single longitudinal cohort at a single center, and generalizability requires external validation across diverse populations, geographic regions, and GCF collection protocols. Differences in laboratory assay platforms, sample collection techniques, and patient populations could affect model transferability.

Third, the observational design cannot establish causal relationships between biomarker trajectories and progression. The identified biomarkers may be correlates rather than drivers of disease progression, and experimental validation is needed to establish mechanistic pathways.

Fourth, the regression and binary classification models were trained separately using the same architectural backbone. Multi-task learning with shared encoder weights, where regression and classification losses are jointly optimized, could potentially improve both tasks through complementary gradient information and merits future investigation.

Finally, participant demographics including age, sex, smoking status, and systemic conditions were not available for subgroup analysis in this secondary analysis. Future studies should examine whether model performance varies across demographic and clinical subgroups, as health equity considerations are central to responsible AI deployment in healthcare.

### 4.7 Future directions

Several promising directions emerge from this work. First, extending the prediction horizon from 2 months to 4- and 6-month windows would provide clinicians with longer lead times for intervention planning. Second, reducing the current 64-protein panel to a smaller core set of the most informative markers would substantially lower assay costs and enable development of targeted chairside point-of-care or home-based tests, providing a molecular supplement to the current clinical periodontal monitoring. Third, extending the framework to predict treatment response would enable personalized therapy selection. Utilizing the GCF monitoring through the treatment phase at 18-month, the temporal model could learn to associate pre-treatment biomarker trajectories with post-treatment outcomes, supporting data-driven decisions about whether a patient is likely to respond to non-surgical therapy alone or requires surgical intervention.

## 5. Conclusions

This study demonstrates that temporal deep learning applied to longitudinal GCF protein profiles enables accurate prediction of clinical parameter changes and reliable classification of both current and future periodontitis progression status. The systematic model development confirmed that explicitly modeling biomarker trajectory dynamics, rather than relying on single-visit snapshots, is the primary driver of prediction performance. Importantly, this framework operates independently of clinical examination data, supporting its potential use in remote monitoring between routine office visits. The identification of a core set of informative biomarkers provides a foundation for developing cost-effective targeted assays for clinical translation.

